# Genomic Evidence for Extra-Intestinal Foodborne Disease due to Zoonotic Bacteria: A Public Health Risk

**DOI:** 10.1101/2024.02.22.24303194

**Authors:** Patrice Landry Koudoum, Luria Leslie Founou, Brice Davy Dimani, Aurelia Mbossi, Jessica Ravanola Zemtsa, Mukanda Gedeon Kadima, Arsène Kabamba-Tshikongo, Jonathan Asante, Karen H. Keddy, Teresa Estrada Garcia, Raspail Carrel Founou

**Affiliations:** Antimicrobial Resistance and Infectious Disease (ARID) Research Unit, Research Institute of Centre of Expertise and Biological Diagnostic of Cameroon (CEDBCAM-RI); Reproductive, Maternal, Newborn and Child Health (ReMARCH) Research Unit, Research Institute of Centre of Expertise and Biological Diagnostic of Cameroon (CEDBCAM-RI); Bioinformatics & Applied Machine Learning Research Unit, EDEN Biosciences Research Institute (EBRI), EDEN Foundation, Yaoundé Cameroon; Antimicrobial Research Unit, School of Health Sciences, College of Health Sciences, University of KwaZulu-Natal, Durban, South Africa; Discipline of Pharmaceutical Sciences, School of Health Sciences, University of KwaZulu-Natal, Durban, South Africa; Clinical Biology Laboratory, Faculty of Pharmaceutical Sciences, Université de Lubumbashi; School of Pharmacy and Pharmaceutical Sciences, University of Cape Coast, Cape Coast, Ghana; Department of Veterinary Tropical Diseases, Faculty of Veterinary Science, University of Pretoria, Pretoria, South Africa; Department of Molecular Biomedicine, CINVESTAV-IPN, Mexico City, Mexico; Department of Microbiology-Haematology and Immunology, Faculty of Medicine and Pharmaceutical Sciences, University of Dschang, Dschang, Cameroon

**Keywords:** Extra-intestinal foodborne disease, antimicrobial resistance, zoonosis, One Health

## Abstract

Foodborne diseases (FBD) constitute a significant worldwide concern with both health and socioeconomic repercussions. FBD predominantly manifest with gastrointestinal symptoms. Nevertheless, extraintestinal infections and symptoms caused by foodborne pathogens are increasingly being reported worldwide. Furthermore, FBD caused by drug resistant bacteria causing both intestinal and extra-intestinal FBD (EIFBD) has further exacerbated their burden. This review (i) summarizes the status of intestinal foodborne pathogens, (ii) delineates the drivers, highlights the (iii) genomic evidence, and (iv) global significance of EIFBD. Understanding the transmission routes and genomic evolution of the principal bacteria causing extra-intestinal FBD is of paramount importance to fully assess the burden of FBD globally, especially in under-resourced regions of Asia, Africa, and Latin America. Acknowledging EIFBD as an integral part of FBD is critical to reach sustainable development. Integrated strategies considering the One Health approach are requisite if FBD are to be controlled, plus to ensure food safety and security.

**Data summary:** The authors confirm that all supporting data, code and protocols have been provided within the article or through supplementary data files.

**Impact Statement:** Foodborne diseases (FBD) present a global health challenge, with the significant health impact and economic costs. Commonly, these illnesses manifest as gastrointestinal troubles; however, there is an alarming increase in reports of foodborne pathogens causing infections beyond the intestinal tract. The complexity of managing FBD is worsened as various pathogens exhibit resistance to antibiotics, a concern applicable to both intestinal and extraintestinal infections.

This review serves several critical functions: (i) provide a current overview of the pathogens responsible for gastrointestinal FBD, (ii) examine the underlying factors contributing to the proliferation of these diseases, (iii) present insights gleaned from genomic studies, which enhance our understanding of these pathogens.

Grasping the mechanisms of pathogen transmission and genetic evolution is essential to accurately evaluate the worldwide impact of FBD. Acknowledging extraintestinal infections as a significant component of FBD is vital for sustainable progress. Combating FBD effectively necessitates a comprehensive “One Health” approach, which integrates the well-being of humans, animals, and the environment. Such a strategy is imperative not only for controlling FBD but also for ensuring the safety and availability of food resources globally.

## INTRODUCTION

Foodborne diseases (FBD) have been defined by the World Health Organization (WHO) as diseases caused by the ingestion of unsafe food contaminated by infectious (including bacteria, viruses, fungi, and parasites) and non-infectious agents (chemicals) (WHO, 2015). In 2010, 600 million FBD cases were recorded with 420,000 deaths (WHO, 2015). FBD are a serious global public health threat that manifest mainly with gastrointestinal symptoms such as nausea, vomiting, and diarrhoea (WHO, 2015). These clinical manifestations of some FBD led to the investigation of foodborne pathogens mainly in the gastrointestinal tract (GIT), whilst some foodborne pathogens can also cause extra-intestinal infections such as urinary tract and bloodstream infections, and meningitis (Manges et al., 2019).

The most common bacteria causing FBD include diarrhoeagenic *Escherichia coli* (DEC) (Riveros et al., 2017; Estrada-Garcia & Navarro-Garcia, 2012), *Salmonella* spp. (Abbott et al., 2012; Jüch et al ., 2013; Pulford et al., 2019), *Listeria monocytogenes* (Schlech, 2019), and *Campylobacter* spp. (Chlebicz & Śliżewska, 2018) have all been acknowledged to cause both intestinal and extra-intestinal manifestations. “Non-classical” foodborne bacteria such as extra-intestinal pathogenic *E. coli* (ExPEC) (Liu et al ., 2018), *Klebsiella pneumoniae* (Davis et al., 2015), Group B *Streptococcus* (GBS) (Boonyayatra et al ., 2020; Simões et al., 2021; Sørensen et al ., 2019) and *Staphylococcus* spp. (Lawal et al., 2021) originating from food and/or food animals have recently been implicated in either human urinary tract infections (UTIs), sepsis, meningitis, or a combination of these diseases, with mild- to life-threatening outcomes. Nevertheless, the scope and burden of extra-intestinal foodborne diseases (EIFBD) caused by these “non-classical” foodborne bacteria are poorly documented worldwide. EIFBD without associated gastrointestinal symptoms are yet to be considered globally, even in the most advanced foodborne disease surveillance systems implemented in high-resource countries which mainly focus on intestinal FBD. This likely leads to an underestimation of the global burden of FBD.

Infectious FBD are frequently due to zoonotic transmission of pathogens from food animals or products of food animals (Gerner-Smidt et al., 2019). Foodborne bacteria cause FBD by numerous ways including, but not limited to the (i) production of toxins in food that is subsequently consumed by humans, (ii) production of toxins in the gastrointestinal tract after ingestion, (iii) colonization of the intestinal tract, and (iv) invasion of intestinal epithelium cells (Antunes et al., 2020). Understanding the different mechanisms and drivers of pathogen emergence, spread, and persistence in populations is essential for designing appropriate prevention and containment measures against infectious diseases (Watkins et al., 2016). The relative abundance of FBD echoes with three main questions: to demonstrate that extraintestinal infections such as UPEC are indeed FBD? Are there genetic evidences of EIFBD globally? Could drug-resistant EIFBD exacerbate the burden of FBD? Therefore, the aims of this review are to: (i) summarize the current trends of intestinal FBD, (ii) delineate the evidence of EIFBD, (iii) describe the genetic evidence of EIFBD, and (iv) outline the global significance of EIFBD, to emphasize data gaps and provide a roadmap for future studies.

## I. Methods

Original articles in French or English which included genetic data characterizing bacterial pathogens associated with extra-intestinal foodborne diseases were searched independently in the MEDLINE database through PUBMED by four authors, PLK, BDD, AM and JRZ. Advanced searches were done using Boolean operators AND, OR, and NOT and the key terms used included: extra-intestinal foodborne disease, foodborne disease, foodborne illness, foodborne urinary tract infections, urinary tract infection, bloodstream infection, bacteraemia, urosepsis, sepsis, meningitis, invasive disease, wound infection, *Enterobacteriaceae*, *Enterobacterales*, *Escherichia coli*, *Klebsiella pneumoniae*, *Staphylococcus*, *Streptococcus pyogenes* (Group A *Streptococcus*, GAS), *Streptococcus agalactiae* (Group B *Streptococcus*, GBS) , genotype, virulence, and molecular epidemiology. All relevant articles, English or French, clearly presenting the genetic link between human infections and animal food and/or food animals, published between 1^st^ January 2015 and 29^th^ April 2022 were included. Reviews, Case reports, and conference proceedings were excluded.

## II. Overview of intestinal foodborne diseases

FBD represent one of the major causes of mortality and morbidity in humans with 600 million cases and 420000 deaths in 2010 alone (WHO, 2015). *Campylobacter* spp*., Salmonella enterica, Shiga-toxin E. coli, Shigella* spp., and *Listeria monocytogenes* respectively are the leading predominant bacteria involved in FBD (Crim et al., 2014). Moreover, several reports have described the role of other bacteria such as *K. pneumoniae, S. aureus* (Ko et al., 2018; Liu et al., 2021), GAS (Karabela et al., 2022), and GBS (Barkham et al., 2018; Tiruvayipati et al., 2021) and that they are transmitted after food consumption, therefore, they should be considered foodborne pathogens. Bacterial contamination can occur at any stage from farm-to-the-plate continuum (Founou et al., 2016; Yang et al., 2017). This section provides a general overview of foodborne bacteria and intestinal foodborne diseases (IFBD).

### 1. Escherichia coli

*Escherichia coli* is a commensal and ubiquitous bacterium that has been involved in numerous cases of intestinal foodborne disease (IFBD) (Hu et al., 2021) mainly through the consumption of contaminated food (Liu et al., 2018). In 2015, the World Health Organization (WHO) published the first estimates of FBD worldwide reporting that *E. coli* caused over 110 million cases of FBD resulting in over 63,000 deaths (WHO, 2015). Diarrhoeagenic *E. coli* (DEC) pathotypes including enteropathogenic (EPEC) and atypical EPEC (lacking the bundle-forming pilus), enterotoxigenic (ETEC), enteroaggregative (EAEC), diffusely adherent (DAEC) entero-invasive (EIEC), and Shiga toxin-producing *E. coli* (STEC), are important causes of diarrhoea in children, of travellers’ diarrhoea (TD), and diarrhoea in immunocompromised patients (Martínez-Santos et al., 2021). STEC can be specifically associated with haemolytic uremic syndrome, while ETEC and EAEC with TD (Cabrera-Sosa & Ochoa, 2020; Jiang & DuPont, 2017).

The foodborne transmission of *E. coli* has been clearly evidenced by its impact on human health (WHO, 2015). A study conducted in South Korea, where the authors performed the molecular characterization of *E. coli* in human stool, environmental and preserved food samples demonstrated that consumption of egg-based foods was associated with atypical EPEC O157:H45 infections with an attack rate of 2.5% (Park et al., 2014).

### 2. Salmonella *spp*

Foodborne *Salmonella* infections contribute substantially to morbidity and mortality worldwide (WHO, 2015). *Salmonella* is of public health concern due to the burden of salmonellosis and frequent reports of AMR. AMR salmonellae are difficult to treat, increasing the number of deaths due to FBD (Chlebicz & Śliżewska, 2018). Misuse of antimicrobials for animal husbandry and in the treatment of infections has impacted the rising incidence of AMR in salmonellosis.

Globally, 3.4 million non-typhoidal *Salmonella* (NTS) infections occur resulting in close to 19,000 deaths yearly (Stanaway et al., 2019). In the United States of America (USA), high-resolution whole genome sequenced (WGS) revealed close phylogenetic relationships between NTS serovars detected in humans, food animals, and agricultural environmental sources different hosts, indicating the importance of pathogens of animal origin in FBD (Pornsukarom et al., 2018). Likewise, another study carried out in China demonstrated with WGS, that S. Typhimurium isolated from pigs was responsible for a large proportion of FBD although this may not be the case in countries where pork is not eaten (Sun et al., 2020).

### 3. Campylobacter spp

Campylobacteriosis is a zoonosis whose main reservoir is the digestive tract of wild or domestic birds, such as poultry, but also other food animals like sheep, cattle, and pigs (Ahmed & Gulhan, 2021). *Campylobacter* species are usually commensal in animals but can spread to humans directly or indirectly through ingestion of contaminated food or water (Facciolà et al., 2017). *Campylobacter* spp. is one of the most important foodborne pathogens even in high-income countries (Facciolà et al., 2017). In 2010, *Campylobacter* spp. caused around 96 million FBD cases and 21,500 deaths worldwide (WHO, 2015). The most common species involved in human infections are *Campylobacter jejuni* and *Campylobacter coli* (Aksomaitiene et al., 2019). *C. jejuni* population structures are weakly clonal, resulting in the plasticity of the *Campylobacter* genome. The latter is susceptible to point mutations and genomic instability and can readily take up foreign DNA and an enormously diverse population irrespective of which typing system is used (Newell et al., 2010). Nevertheless, some stable clones of *C. jejuni* have persisted, and such clones may provide insight on changing trends (Newell et al., 2010).

By genotypically characterising with multi-locus sequence typing (MLST) *C. jejuni* isolates in different sources of isolation (human, chicken, dairy cattle, and wild birds), in Lithuania, Aksomaitiene et al. (2019) showed that from a total of 341 isolates originating from humans, chickens, cattle and wild birds, CC1034 and CC403 sequence types (ST) were found in all hosts demonstrating that cattle and wild birds were the main reservoirs of *C. jejuni* isolated from human infections (Aksomaitiene et al., 2019).

### 4. Staphylococcus aureus

Staphylococcal food poisoning is one of the most common FBD in the world. Approximately 241,000 cases of FBD are caused by *S. aureus* every year in the USA (Scallan et al., 2011). Raw or undercooked food, especially food of animal origin and nasal carriage by food workers are identified as important reservoirs of *S. aureus* causing FBD (Wang et al., 2017). The ingestion of staphylococcal toxins produced by certain strains is the main cause of disease symptoms (Hennekinne et al., 2012). Some virulence genes such as those encoding for the toxic shock syndrome toxin 1 (TSST-1), exfoliative toxins, and cytolytic toxins are involved in staphylococcal food poisoning (Argudín et al., 2010). In an epidemiological surveillance study in China, 163 *S. aureus* isolates were analysed by pulse field gel electrophoresis (PFGE) with some isolates originating from foodstuffs such as rice noodles and rice rolls, showing identical profiles with those collected in humans with clinical signs of food poisoning (Liao et al., 2018). Likewise, Lui et al. (2021) performed a genotypic characterisation of *S. aureus* isolated from animal samples collected at farms and markets in Xinjiang province and demonstrated that meat originating from abattoirs and farms was contaminated with virulent *S. aureus*. They confirmed the spread of these virulent lineages of *S. aureus* in the food chain and their involvement in FBD (Liu et al., 2021).

## III. Drivers of extra-intestinal foodborne diseases

A complication of diarrhoeal foodborne diseases is their capacity, in some cases, to become systemic diseases (Hoffmann et al., 2020). Intestinal barriers existing prevent any harmful elements including toxins and bacteria from disseminating to the bloodstream and other organs or tissues (Li & Tan, 2020). Factors that directly or indirectly allow foodborne bacteria to breach this barrier are EIFBD drivers. The drivers of EIFBD include a triad of (i) host susceptibility (ii) genomic diversity within pathogens, and (iii) food contamination, which are known to drive the transmission of pathogenic bacteria to humans as seen in figure 1 below (Antunes et al., 2020). While food contamination can directly or indirectly lead to EIFBD, bacterial pathogenicity, and host susceptibility are favouring factors which might lead to an EIFBD after an IFBD has already been established (Antunes et al., 2020).

**Figure 1.**
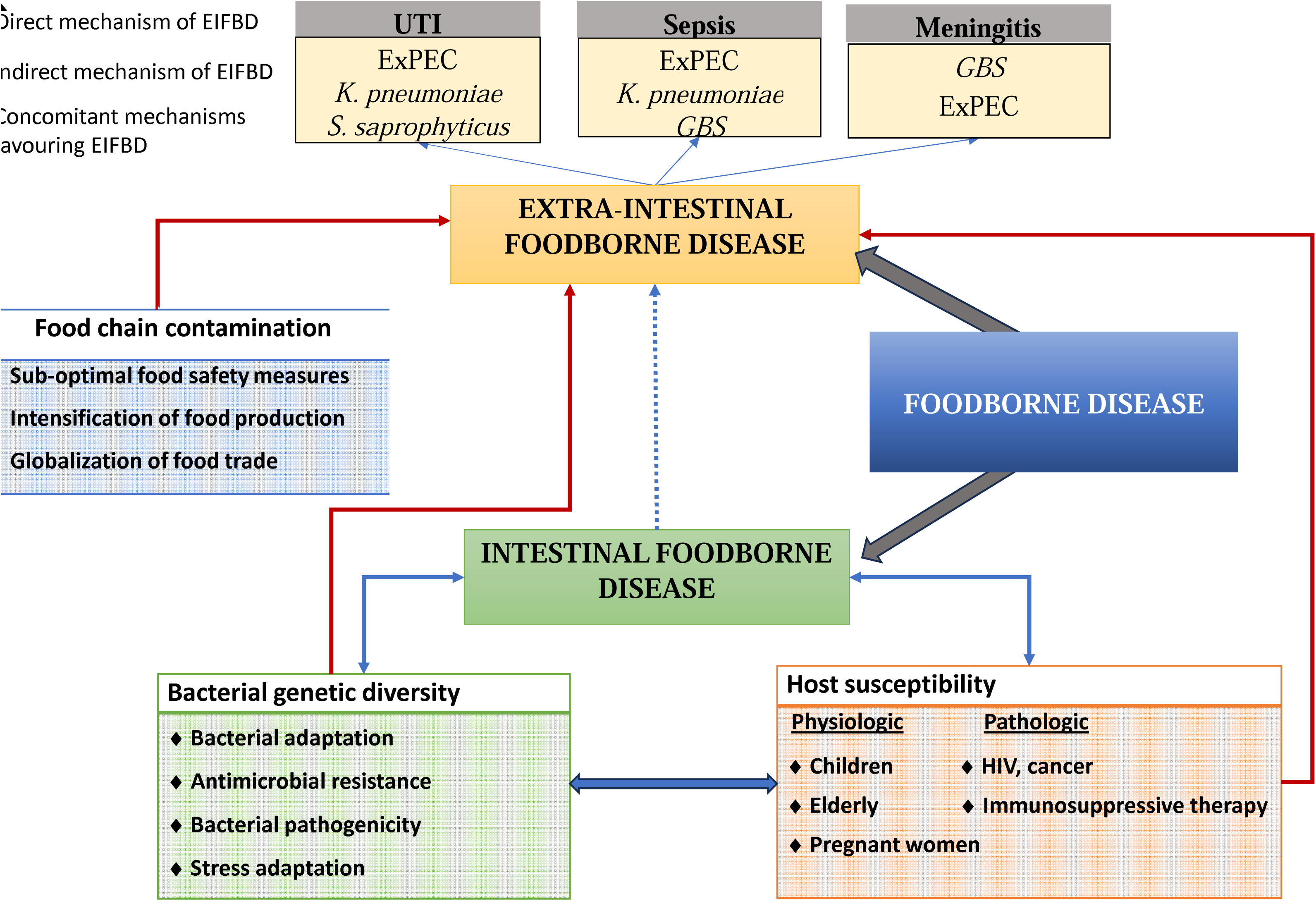
Drivers of extra-intestinal foodborne diseases.

### 1. Host susceptibility

The main host factor favouring EIFBD is immunosuppression, which increases the host’s vulnerability to pathogens (Lund & O’Brien, 2011). One of the main vulnerable groups to infections due to FBP are undernourished children from sub-Saharan Africa and Latin America, their increased susceptibility to diarrhoeagenic pathogens has clearly been established (Acosta et al., 2016; Opintan et al., 2010).

Other vulnerable groups include people living with acquired immunodeficiency syndrome (AIDS), people with physiologically impaired immune systems like pregnant women, the elderly as well as individuals on immune suppressive drugs or cancer therapy (Lund & O’Brien, 2011). This vulnerability leads to a reduction of the load of organisms required to cause infectious diseases and, is therefore, easily associated with bacterial dissemination to extra-intestinal sites due to the lowering of host’s immune defences which are supposed to prevent this (Lund & O’Brien, 2011). Immunocompromised individuals are thus more prone to have an EIFBD and more severe diseases caused by extra-intestinal dissemination.

### 2. Foodborne bacteria pathogenicity and diversity

Bacteria are subjected to adverse conditions that contribute to their stress, favouring their survival, or acquisition of new traits (Begley & Hill, 2015). These traits are responsible for virulence in foodborne pathogens and differ between species and subtypes (Table 1). The acquisition of virulence factors involved in pathogenicity and of resistance genes leading to antimicrobial resistance or exposure to a hostile environment may allow bacteria to become more virulent, persist in the host by counteracting the innate and adaptive immune responses, spread to other tissues, and subsequently lead to extra-intestinal infections (Cavestri, 2020).

**Table 1.** Virulence factors of the major bacterial pathogens associated with foodborne disease.

| Foodborne pathogen | Virulence factor | Mechanisms of action | References |
| --- | --- | --- | --- |
| EAEC | *aggR* | Transcriptional activator, regulating the expression of various virulence factors encoded by the chromosome and pAA plasmid | (Pakbin, et al., 2021) |
|  | Aggregative Adherence Fimbriae (*aggA*, *aafA*, *agg3A*, *agg4A*, *agg5A*) | Mediate attachment to the intestinal mucosa and trigger host's inflammatory response. They also promote biofilm formation. | (Boll et al., 2017; Moraes et al., 2020; Moran-Garcia et al., 2022) |
|  | Dispersin (*aap*) | Overcomes the electrostatic attraction between fimbriae and the bacterial surface to ease the dispersion of EAEC across the intestinal mucosa. | (Moraes et al., 2020) |
|  | pAA | Plasmid that encodes for AAFs and toxins such as the autotransporter Pet and the heat-stable enterotoxin *astA*. | (Jønsson et al., 2017) |
| ETEC | Colonisation factors (CFA/I, CFA/II, CFA/IV) | Allow ETEC strains to attach the proximal small intestine enterocytes. | (Vidal et al., 2019) |
|  | Labile toxin (LT I, LT II) | Activate the production of cAMP leading to the activation of cellular kinases in charge of modulating the activity of sodium and chloride channels in the apical membrane of intestinal epithelial cells resulting in the efflux of salt and water in the intestinal lumen, and hence a watery diarrhoea. | (Sheikh et al., 2022) |
|  | Heat-stable toxins (STa, STb) | Binds its receptor sulfatide, on the surface of intestinal epithelial cells resulting in the activation of a pertussis toxin-sensitive GTP-binding regulatory protein. This leads to an influx of calcium into the cell and activation of calmodulin dependent protein kinase II which in turns opens Calcium-activated Chloride Channel (CaCC) allowing for the secretion of chloride into the intestinal lumen, resulting in watery diarrhoea. | (Sheikh et al., 2022) |
| EPEC | Type IV bundle-forming pilus (*bfp*) | Formation of bacteria autoaggregate and localised adherence. | (Pakbin et al., 2021) |
|  | *eae* | Permits the attaching and effacing of enterocytes. | (Pakbin et al., 2021) |
| EHEC | Shigatoxin (*stx1* and *stx2*) | Enters cells by endocytosis and is transported to the Golgi apparatus and then to the endoplasmic reticulum where it targets ribosomes, inactivating it and thus inhibiting protein synthesis. | (Pacheco & Sperandio, 2012) |
| Salmonella spp. | *Salmonella* plasmid virulence (*spvB*) | Plasmid contributing to the loss of intestinal barrier integrity and promotes extra-intestinal dissemination of infection by facilitating translocation of *Salmonella*. The *spvB* gene is commonly indexed in the systemic dissemination of *Salmonella* infection and secreted into host cells via *Salmonella* pathogenicity islands (SPI-2) T3SS | (Sun et al., 2020) |

EAEC: Enteraggregative *Escherichia coli*; ETEC: Enterotoxigenic *Escherichia coli*; EPEC: Enteropathogenic *Escherichia coli*, EHEC: Enterohemorrhagic *Escherichia coli*

The presence in the human host of foodborne bacteria harbouring antibiotic-resistance genes could favour extra-intestinal dissemination of these pathogens because antibiotics do not eliminate them, and microorganisms could then colonize other niches. For instance, when the host’s immune system fails to eradicate antimicrobial-resistant bacteria from the gastrointestinal tract, bacteria might persist within the gut for an extended duration. Consequently, they could breach the intestinal barrier and enter the systemic circulation, potentially resulting in an extraintestinal infection. This is supported by the existence of virulence genes co-located along with antibiotic resistance genes encoded on mobile genetic elements such as plasmids (Beceiro et al., 2013; Singer, 2015). For instance, non-typhoidal *Salmonella* (NTS) isolated from retail meat were involved in infections in children and those infected with drug-resistant NTS strains were more likely to develop severe sepsis than those with the drug-susceptible *Salmonella* (Chang et al., 2020).

### 3. Food contamination

In recent years the emergence, dissemination, persistence, and exposure of highly resistant/virulent foodborne bacteria at each step of the food production chain has increased due to the intensification of food production activities in the agricultural and livestock sectors, and large-scale distribution of food products to cope with the global food demand (Founou et al., 2021). This is particularly true in low- and middle-income countries (LMICs) as a result of poor hygiene and sanitation, inadequate handling processes, insufficient safety control measures during operations, poor control of the use of antibiotics as growth promoters, and sub-optimal storage facilities (Founou et al., 2021). Food contamination can occur through direct spread from infected animals or animal excretions, or via contamination with foodborne bacteria on the hands of food handlers (Founou et al., 2021; Founou et al., 2016). Although gastrointestinal disease is usually self-limiting, systemic dissemination may occur if vulnerable patients infected with resistant pathogens are treated with the incorrect antibiotic. The globalization of trade of food products is a major factor contributing to the spread of foodborne pathogens internationally (Tiseo et al., 2020) and AMR pathogens causing both IFBD and EIFBD can cross country/continental borders when contaminated food products are transported from one region of the world to another.

## IV. Burden and genetic evidence of extra-intestinal foodborne diseases

Foodborne bacteria are now increasingly being recognised in extra-intestinal infections (Riley, 2020), but because of (i) the relatively long incubation period between gastro-intestinal tract (GIT) colonization and dissemination to other body systems, (ii) poor understanding of this paradigm by physicians, and (iii) insufficient investigation of the transmission routes associated with invasive infections, it is difficult to ascertain causality in most cases. Nevertheless, some studies provided genetic evidence on EIFBD especially urinary tract infections (UTIs), sepsis, and meningitis, that were caused by *E. coli*, *K. pneumonia*, *S. saprophyticus*, and Group B *Streptococcus* (GBS) as disclosed below and in Table 2.

**Table 2:** Summary of the genetic evidence of extra-intestinal foodborne bacterial diseases.

| Country | Study period | Study Objective | Bacteria | Human infection | Food/Animal Source | Genetic techniques | Main findings | Authors |
| --- | --- | --- | --- | --- | --- | --- | --- | --- |
| USA | 2011-2012 | Assess the genetic relatedness, virulence and antibiotic resistance rates between *K. pneumoniae* isolates from UTIs, BSI and retail meat samples. | *K. pneumoniae* | UTI<br>BSI | Pork, turkey, chicken | MLST, WGS and SNP phylogeny | Four identical MLST pairs were found in both clinical and retail meat samples. Following phylogenetic analysis these pairs were closely related and displayed similar virulence profile using murine sepsis model. | (Davis et al., 2015) |
| USA | January-December 2012 | Evaluate the role of retail meat as a potential source of *ST131-H22* causing human UTI and BSI. | *E. coli ST131-H22* | UTI<br>BSI | Pork, chicken | WGS and SNP-based phylogeny | Six of human clinical *E. coli ST131-H22* strains were found in the same clade with meat and were closely related. Clinical strains harbouring CoIV plasmid, a plasmid ascribed APEC were more related to meat strains than those without the plasmid | (Liu et al., 2018) |
| Worldwide | 1997-2017 | Investigate the foodborne origin and spread of *S. saprophyticus* causing human UTI | *S. saprophyticus* | UTI | Pig, pork | WGS and SNP-based phylogeny | The meat food chain was an important source of *S. saprophyticus* causing human UTI as the live pig, meat and UTI *S. saprophyticus* strains were closely related after phylogenetic analysis. | (Lawal et al., 2021) |
| Brazil | 2019 | Assess the phylogenetic link between bovine milk GBS isolates and those from human invasive disease | GBS | Sepsis<br>Meningitis | Bovine milk | MLST, PFGE | Two blood, one CSF, and one bovine milk GBS serotype III/ST17 strains were highly related to each other ($\geq 80\%$ similarity). These strains also had identical resistance profiles, harboured the same resistance and virulence genes tested. | (Simões et al., 2021) |
| Thailand<br>Vietnam | 2008-2016 | Perform the comparative genetic analysis of GBS strains from diseased fishes in Thailand and Vietnam | GBS | Meningitis | Fish | WGS and cgMLST | Thailand GBS ST283 strains were closely related to the Singaporean strain causing foodborne human invasive disease in 2015 | (Kayansamruaj et al., 2019) |
| Thailand | 2011-2014 | Evaluate the genetic relatedness between GBS isolates from human sepsis, bovine mastitis and from fishes | GBS | Sepsis | Cattle, fish | MLST and PFGE | One human blood GBS serotype III was closely related ($\geq 80\%$ similarity) to five identical (100% similarity) fish GBS serotype III isolates. A human blood sample was closely related to a bovine milk isolate. | (Boonyayatra et al., 2020) |

### 1. Urinary tract infection (UTI)

Urinary tract infections (UTIs) are a serious public health problem and are the most common bacterial infections in women of economically active age worldwide with over 150 million cases yearly (Flores-Mireles et al., 2015). Foodborne bacteria can transiently colonize the human gastrointestinal tract after ingestion (either directly after contact with an infected/contaminated animal or indirectly via food or contaminated surfaces) where they could later spread to the urinary tract (Flores-Mireles et al., 2015). This hypothesis is more plausible and thus more beneficial for bacteria invading the female urogenital tract since women have a shorter urethra than men (Nordstrom et al., 2013). Bacteria can ascend from the lower urinary tract to the ureter and then to the kidneys, causing pyelonephritis. From the kidneys, bacteria can disseminate to the bloodstream, known as urosepsis, a life-threatening condition with high mortality rates of 20-40% (Dreger et al., 2015; Riley, 2020).

#### 1.1. Escherichia coli

Uropathogenic *E. coli* (UPEC) is the single most prevalent uropathogen causing 70-95% of all UTIs (Singer, 2015) and belonging to the broad group of extra-intestinal pathogenic *E. coli* (ExPEC) having virulence factors enable it to thrive in environments other than the human gastro-intestinal tract including the human urogenital tract (Flores-Mireles et al., 2015). Liu et *al.* prospectively assessed the role of foodborne *E. coli* ST131-H22 as an uropathogen using core genome single nucleotide polymorphisms (cgSNP)-based phylogenetic analysis (Liu et al., 2018). *E. coli* ST131 detected in retail meat (pork and chicken) were closely related to *E. coli* isolates from human clinical samples including urine and blood (Liu et al., 2018). Moreover, human clinical *E. coli* ST131-H22 strains harbouring the CoIV plasmid, typically ascribed to avian pathogenic *E. coli*, were more closely related to *E. coli* ST131-H22 strains from meat sources, in comparison to those strains lacking the ColV plasmid. This suggests that poultry could be a source of not only intestinal but also extra-intestinal human infections such as UTI and urosepsis (Liu et al., 2018).

#### 1.2. Klebsiella pneumoniae

Though not acknowledged as a “classical” foodborne pathogen, *K. pneumoniae* strains originating from animals have also been identified in clinical infections. Davis et al., (2015), reported that *K. pneumoniae* strains, isolated from urine samples of UTI patients, exhibited a high degree of genetic similarity to strains found in retail meat in the USA (Davis et al., 2015). The authors analysed 508 retail meat samples (pork, turkey, and chicken) and 1728 clinical urine samples from UTIs patients, and the core genome SNP-based phylogenetic analysis of 83 *K. pneumoniae* isolates from both retail meat and urine samples, revealed five closely related human-meat isolate pairs were genomically closely related (Davis et al., 2015). Furthermore, Davis et al. (2015) study revealed that after testing in a murine sepsis model, the five *K. pneumoniae* isolate pairs displayed a comparable virulence profile. The authors concluded that the high genetic relatedness and similar virulence profile strongly suggested that UTI-causing *K. pneumoniae* could likely have an animal origin, emphasizing the need for further consideration of *K. pneumoniae* in EIFBD (Davis et al., 2015).

#### 1.3. Staphyloccus saprophyticus

*S. saprophyticus* is the one of most prevalent Gram-positive uropathogens causing uncomplicated UTIs (Flores-Mireles et al., 2015). De Paiva-Santos et *al*. after analyzing the virulome of 142 *S. saprophyticus* isolates collected from cheese, recreational water, and urine specimens of UTI patients found similar virulence gene profiles among the strains from all sources (de Paiva-Santos et al., 2018). Based on these results, the authors surmised the elevated uropathogenic potential of foodborne (cheese-borne) *S. saprophyticus* isolates (de Paiva-Santos et al., 2018). Likewise, SNP-based phylogenetic analysis of 128 *S. saprophyticus* isolates causing UTIs and 104 isolates from several sources of the slaughterhouse including from pigs, raw pork meat, and the slaughterhouse environment, workers, and equipment demonstrated that *S. saprophyticus* causing UTIs was of foodborne origin since the *S. saprophyticus* isolates from the pig and raw meat clustered together with the UTI patient, and slaughterhouse workers *S. saprophyticus* isolates (Lawal et al., 2021). Interestingly, all *S. saprophyticus* isolates from this cluster harboured the antiseptic and disinfectant-resistant gene *qacA*, its expression favours the persistence of these bacteria on hands and contaminated surfaces even after disinfection, thus enabling *S. saprophyticus* transmission at the animal-environment-human interface (Lawal et al., 2021).

### 2. Sepsis and bloodstream infections

Group B *Streptococcus* (GBS) has gained notoriety as the leading cause of neonatal infections and deaths with high mortality rates globally (Seale et al., 2017). Group B *Streptococcus* has been associated with cattle and more recently with fish and some lineages are considered as foodborne bacteria (Kayansamruaj et al., 2019). Furthermore, some studies have reported indistinguishable and highly related PFGE profiles between bovine, fish, and human GBS isolates (Boonyayatra et al., 2020; Simões et al., 2021; Sørensen et al., 2019). For instance, Simões et *al.* (2021) found a closely related hypervirulent GBS serotype III/ST17 between isolates from bovine milk and that from a neonate presenting with GBS infection albeit the authors did not specify if the mother was screened for carriage (Simões et al., 2021). Of interest, these GBS isolates not only shared identical MLSTs and similar PFGE profiles but (i) had identical antimicrobial resistance profiles, (ii) harboured the hyper-virulent GBS adhesin (HvgA) *gbs2018-C* gene, genes encoding for pilus variants PI-1, PI-2a and PI-2b, (iii) possessed DNase activity and (iv) harboured the *tet*M gene (Simões et al., 2021). The fact that these GBS strains have similar phenotypic, genotypic, and high-resolution phylogenetic characteristics strongly underscores the role of bovine GBS serotype III/ST17 in the vulnerable neonatal and infant population.

### 3. Meningitis

Bacterial meningitis is still a global health problem, associated with high morbidity and mortality worldwide (Bystritsky & Chow, 2022). Considerable progresses have already been made in the diagnosis and treatment of bacterial meningitis caused by frequently incriminated pathogens such as *S. pneumoniae*, *N. meningitidis*, and GBS (van de Beek et al., 2021). GBS is among the leading cause of meningitis in newborns who inhale or ingest amniotic fluid before or during delivery (Fischer et al., 2021). Of the ten known GBS capsular serotypes, six (Ia, Ib, II, III, IV, and V) are the most frequent serotypes associated with invasive disease and the clone GBS III/ST17, associated with GBS meningitis (Pellegrini et al., 2022). Kayansamruaj et *al.* (2019). In a longitudinal study conducted in Thailand and Vietnam the comparison between 21 sequenced GBS genomes by phylogenetic analysis revealed a correlation between the genomes of species isolated from fish and those causing neonatal meningitis in both countries (Kayansamruaj et al., 2019). Barkham et al (Barkham et al., 2019) reported that GBS ST283 strains isolated from diseased fishes in Thailand were closely related to the GBS ST28 strain causing human invasive disease in Singapore (Kayansamruaj et al., 2019). The above-mentioned studies strongly underline the role of both bovines and fishes as reservoirs of virulent GBS strains and their contribution to human invasive disease. They further suggest that a subset of the meningitis cases occurring worldwide might likely have a foodborne origin.

## V. Antimicrobial resistance and extra-intestinal foodborne diseases

Antimicrobial resistance (AMR) is one of the greatest health threats of the 21^st^ century. According to the Global Research on Antimicrobial Resistance Project (*GRAM*) report, antibiotic-resistant bacterial infections were associated with nearly 5 million deaths and were directly responsible for 1.27 million deaths globally, in 2019 (Murray et al., 2022). The global increasing rate of AMR is alarming, and the world is making a day-to-day step into a post-antibiotic era. The use of antimicrobials in food animal production as growth promoters, for metaphylactic or prophylactic purposes is further exacerbating this concern (Tiseo et al., 2020). Moreover, the global use of antibiotics in food animal production in 2017 was estimated to be 93,309 tonnes and it is expected to rise to 104,079 tonnes by 2030 (Tiseo et al., 2020). More worrisome is that 73% of all sales of antibiotics worldwide including those of clinical importance are for use in the food producing industry (Van Boeckel et al., 2019). Antibiotic use in food-animal production is reportedly known to trigger a selection pressure leading to the rapid emergence and spread of antibiotic-resistant bacteria (ARB) and antibiotic-resistant genes (ARGs) at the animal-human-environment interfaces (Founou et al., 2021). AMR is a quintessential One Health issue needing urgent global and multifaceted actions for its containment.

It is well known that AMR reduces therapeutic choices, thus causing difficult-to-treat infections or increasing mortality rates (Murray et al., 2022). There is a reported increase in the prevalence of antimicrobial resistance in virulent *E. coli*, especially *E. coli* ST131 ExPEC strains isolated from food animals and humans (Singer, 2015). The study conducted by Liu et *al.* (2018) on *E. coli* ST131-H22 previously discussed above showed a high level of multidrug resistance (80% for animal sources and 33% for human sources), with some isolates harbouring mobile colistin resistance (*mcr*) gene (Liu et al., 2018). Resistance to the last resort antibiotic drug, colistin, is of particular concern as resistant bacteria can disseminate from animals to humans (Liu et al., 2018), resulting in difficult-to-treat extra-intestinal diseases with restricted treatment options and associated higher mortality rates.

## VI. The global significance of extra-intestinal foodborne diseases

EIFBD are important threats that should be considered as part of FBD surveillance systems. The pathogen that typically displays the significance of EIFBD are ExPEC group, which causes numerous human health infections globally with both economic and social costs (Manges et al., 2019). Nevertheless, the clinical and socioeconomic impact of ExPEC infections, especially regarding increasing AMR, is underappreciated and neglected (Manges et al., 2019). Extraintestinal pathogenic *Escherichia coli* have been linked with several environmental reservoirs and transmission routes. One of the major challenges in recognising EIFBD and ExPEC infections as a public health threat is the absence of routine molecular investigation or genomics studies (Manges et al., 2019). In the absence of large-scale, routine, public health reporting, or surveillance systems that consider the One Health compartments, it is not possible to capture information on circulating lineages nor to provide a complete snapshot of the genomic evolution, transmission dynamics, and lineages of ExPEC and other pathogens causing EIFBD.

Most of the large-scale genomic studies of ExPEC have been conducted in high income countries of Europe and North America (Manges et al., 2019) revealing the necessity of conducting more studies worldwide, particularly in low-and middle-income countries (LMICs). Thus, data from Asia, Africa and Latin America are scarce despite recent efforts to improve regional capacity to perform genomic analyses, but they are still very expensive for these countries. Understanding the genomic epidemiology of key circulating lineages in these regions is important in order to decipher the transmission routes of foodborne pathogens, predict their likely public health implication, to implement sustainable infection prevention and control (IPC) measures nationally, regionally, and internationally. The importance of these studies was clearly shown by a study conducted in Yucatan, Mexico, which revealed a phenotypic (identical drug resistance profiles) and genetical relatedness by PFGE between NTS isolated from human clinical cases, asymptomatic children of Salmonella, suggesting that dissemination through the food chain occurs (Zaidi et al., 2006).

Another important challenge to recognize the significance of EIFBD is the distinction between commensal and pathogenic lineages (Riley, 2020). The high resolution provided by whole-genome sequencing can allow the detection and appreciation of the wide variety of extra-intestinal illnesses caused by foodborne bacteria. However, even when genomic data are available, it is still challenging to clearly define some ExPEC strains and estimate their disease burden as there are no exclusive ExPEC-defining genes as most of these genes are shared with commensal lineages (Sarowska et al., 2019). The global food production and distribution systems coupled with the increasing antimicrobial resistance likely contribute to higher incidence and associated mortality due to EIFBD (Riley, 2020). In this era marked by globalization of trade and humans as well as global population growth, it is important to acknowledge extra-intestinal foodborne bacteria including ExPEC as overlooked “killers”, yet potentially important endemic issues especially in LMICs (Russo & Johnson, 2003).

Additionally, the role of certain bacterial lineages in the global spread of AMR and antibiotic resistance genes has not been fully established, nor has the overall contribution of resistant EIFBD to the global burden of FBD. Extraintestinal foodborne diseases have traditionally not retained a level of attention commensurate with their contribution to morbidity, mortality, and socioeconomic costs in the past, likely because causative bacteria were highly antibiotic susceptible, consequently, easily eradicated with first-line antibiotic therapy (Ventola, 2015). Unfortunately, the global dissemination of antibiotic-resistant and of highly virulent pathogens has recently changed this paradigm. A retrospective study performed in the USA assessing AMR in *E. coli* isolated from humans and food animals including pigs, chickens, and cattle revealed an exponential increase in drug resistance over a 42 years-period (1950-2002) (Tadesse et al., 2012). Out of the 1,729 *E. coli* isolates tested against 15 antibiotics during this period, a statistically significant (<0.001) upward trend in resistance was seen for ampicillin, sulfamethoxazole, and tetracycline (Tadesse et al., 2012). Likewise, animal strains displayed increased resistance to 11/15 antibiotics including gentamicin, sulphonamide, and ampicillin. The authors further revealed that the level of multidrug resistance (resistance to three or more classes of antibiotics) increased from 7.2% in 1950 to 63.6% during the 2000s (Tadesse et al., 2012). Furthermore, strains from the ExPEC group have the notorious distinction of harbouring new emerging antibiotic resistance genes (Manges et al., 2019). The WHO estimated that the global burden of FBD due to foodborne bacteria, viruses, and protozoa, particularly NTS and EPEC, caused nearly 600 million cases of infections, 200,000 cases of deaths, and these were attributed to 16 million disability adjusted life years (DALYs) in 2010 (WHO, 2015). This burden would be exacerbated by the inclusion of EIFBD. Hundreds of thousands of lives, millions of workdays, and billions of healthcare dollars, are lost every year due to EIFBD (Russo & Johnson, 2003). New treatment and prevention measures for EIFBD are needed to improve outcomes whilst reducing global EIFBD burdens. Vaccine development and deployment strategies targeting foodborne bacteria could also be suboptimal if EIFBD are not considered in population structure and transmission dynamic studies.

## Conclusion

Bacterial FBD is a global public health issue with multiple clinical and socioeconomic implications, including escalating AMR. This review focuses on the contribution of EIFBD to the global burden of FBD. It argues that recognizing EIFBD as an integral part of FBD is critical to assessing the true burden of FBD. It also emphasized the instrumental role of genomic and phenotypic (drug-resistance profiles) studies, since several studies have shown genetic relatedness is accompanied by similar drug resistance profiles. Therefore, together the genotypic and phenotypic characterization of isolates from clinical cases and food sources will be pivotal to understanding, detecting, preventing, and containing FBD, and in designing tailored interventions. It further underscores that identifying the transmission routes, the genomic structure, the virulence profiles, and the evolution of the principal circulating lineages, especially in underrepresented regions of Asia and Africa, is of paramount importance to fully assess the contribution of EIFBD to the global burden of FBD. An integrated strategy considering the One Health approach is requisite if we are to address FBD and ensure food safety and security globally.

## Data Availability

The authors confirm that all supporting data, code and protocols have been provided within the article or through supplementary data files.

## Author contributions

**PLK** co-conceptualized the study, searched the literature, extracted, and collated data, and drafted the manuscript. **DBD**, **AM** and **JRZ**, searched the literature, extracted, and collated data, and drafted the manuscript. **LLF** co-conceptualized the study, vetted the data, supervised and drafted the manuscript. **MGK, AKT, JA, KHK, TEG,** and **RCF** provided substantial contribution in writing and revising the manuscript. All authors read and approved the final manuscript.

### Transparency declaration

The authors declare that there is no conflict that could be construed as a financial interest.

### Funding

This research received no specific grant from any funding agency in the public, commercial or not-for-profit sectors.

## Notes

### Competing Interest Statement

The authors have declared no competing interest.

### Funding Statement

The study was funded by the Thrasher Research Fund through the Thrasher Early Career Award awarded to FLL (Award number 01364). The funder had no role in the study design, preparation of the manuscript nor the decision to submit the work for publication.

